# Disparities in COVID-19 Related Mortality in U.S. Prisons and the General Population

**DOI:** 10.1101/2020.09.17.20183392

**Authors:** Kathryn M. Nowotny, David Cloud, Alysse G. Wurcel, Lauren Brinkley-Rubinstein

## Abstract

We provide an analysis of COVID-19 mortality data to assess the potential magnitude of COVID-19 among prison residents. Data were pooled from Covid Prison Project and multiple publicly available national and state level sources. Data analyses consisted of standard epidemiologic and demographic estimates. A single case study was included to generate a more in-depth and multi-faceted understanding of COVID-19 mortality in prisons. The increase in crude COVID-19 mortality rates for the prison population has outpaced the rates for the general population. People in prison experienced a significantly higher mortality burden compared to the general population (standardized mortality ratio (SMR) = 2.75; 95% confidence interval = 2.54, 2.96). For a handful of states (n = 5), these disparities were more extreme, with SMRs ranging from 5.55 to 10.56. Four states reported COVID-19 related death counts that are more than 50% of expected deaths from all-causes in a calendar year. The case study suggested there was also variation in mortality among units within prison systems, with geriatric facilities potentially at highest risk. Understanding the dynamic trends in COVID-19 mortality in prisons as they move in and out of “hotspot” status is critical.

---

Forecasting reported by the Centers for Disease Control and Prevention suggests that there will likely be between 150,000 and 170,000 COVID-19 related deaths in the U.S. by August 8, 2020^1^. The United States ranks 8^th^ in per capita deaths globally^2^. Facilities such as nursing homes, correctional facilities, meat packing plants, and cruise ships have reported large single site outbreaks, with nursing homes accounting for about one-quarter of all deaths^3^. Prisons and jails, however, account for 39 of the 50 largest COVID-19 outbreaks in the United States^4^.

In response to these outbreaks and calls for increased transparency, many correctional systems, federal and state prisons in particular, are now publicly reporting data on COVID-19 testing, confirmed cases, and deaths among incarcerated people and staff. As prisons represent the epicenter for continued outbreaks, an understanding of the disproportionate impact of mortality in prisons is crucial to improving public health and preventing deaths. We provide an analysis of COVID-19 mortality data to assess the potential magnitude of COVID-19 among prison residents and to contextualize COVID-19 deaths in prisons.

## Methods

The primary data for this study are from the COVID Prison Project (CPP) (www.covidprisonproject.com). The CPP publishes an aggregate dataset examining COVID-19 in correctional facilities, including data on the number of tests, the number of confirmed positive cases, and mortality due to COVID-19 among correctional staff and incarcerated individuals. CPP tracks COVID-19 data for 53 prison systems including the 50 states, Puerto Rico, the Federal Bureau of Prisons (BOP), and Immigration and Customs Enforcement (ICE) detention centers, from April 22, 2020 to the present. Only prison systems reporting at least one COVID-19 death as of July 15 2020 were included in the present analysis. Data from ICE, which CPP started collecting after April 22, were excluded. The states not reporting COVID-19 mortality data that were excluded from the analysis include AK, AR, ID, ME, MT, NE, NV, NH, OK, VT, WV, WI, WV, and WY. Additional national data were pooled from a variety of sources including COVID Tracking Project^5^, Centers for Disease Control and Prevention^6^, American Community Survey^7,8^, Bureau of Justice Statistics^9,10^, Vera Institute of Justice^11^. For each source, we use the latest year that data is publicly available. Data analyses consisted of standard epidemiologic and demographic estimates. This research is exempt from Human Subjects review.

We included Texas as a single case study to generate a more in-depth and multi-faceted understanding of COVID-19 mortality in prisons. Texas was chosen for this case study because of its relatively robust reporting of COVID-19 deaths in prison. Additional data for the case study came from Texas Department of Criminal Justice^12–14^, Texas Department of State Health Services^15^, Texas Population Estimates Program^16^, and The Texas Tribune^17^.

## Results

On April 22, there were 97 COVID-19 related deaths among residents reported by U.S. prison systems. Over the course of 3 months, the number of deaths among people confined to U.S. prisons increased to 683 on July 15, a roughly 600% increase or an average of 48 deaths per week. Figure 1 shows the growth in crude COVID-19 mortality rates for the prison and general population. Beginning in early May, the COVID-19 death rate in prisons began to outpace the general population rate. This is reflected in the weekly standardized mortality ratios (SMR), following the CDC data reporting schedule^6^. Adjusting for age and sex, the SMR was not significant on April 25. Beginning the week of May 2, the SMR was 1.59, meaning that adjusting for age and sex the prison population had a COVID-19 mortality rate 159% higher than the general population. By July 11, the SMR was 2.75. On July 15, the latest date that daily data were available, the crude prison COVID-19 mortality rate exceeded the rate for the general population: 50 per 100,000 compared to 40 per 100,000, respectively.

**Figure 1.**
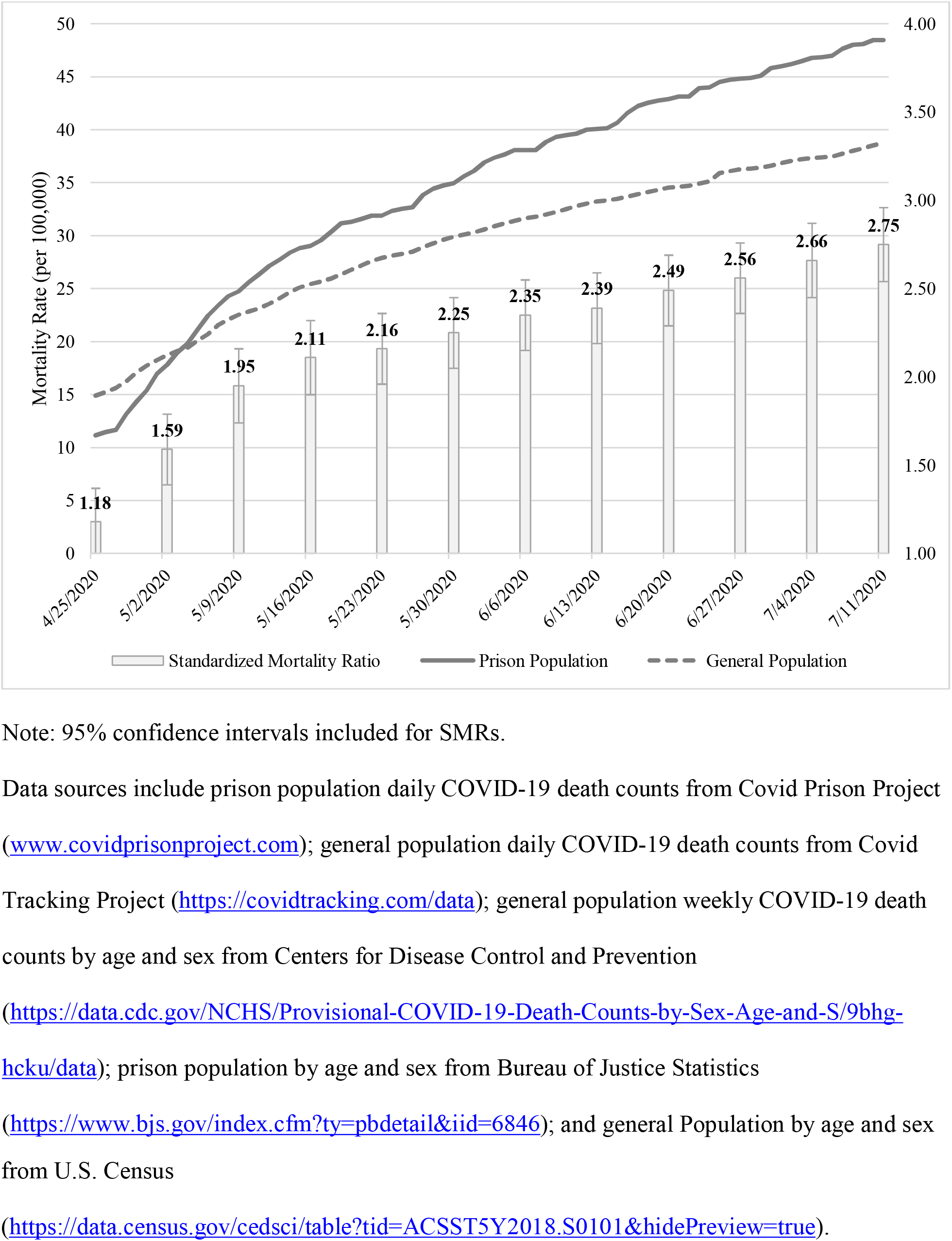
Daily Crude Mortality Rates (per 100,000) for COVID-19 Related Deaths and Weekly Standardized Mortality Ratios (SMR) over Time for the Prison Population and General Population, April 25 to July 11, 2020

Figure 2 shows that there was variation in the relationship between prison and general population mortality across states. Thirty-two out of 50 state departments of correction had reported at least one COVID-19 related death. Among those states, 10 reported little to no difference in mortality between the prison and general population (WA to FL), six states reported a slightly higher mortality rate in the general population (MA to CO), and six states reported a substantively higher mortality rate in the general population (NY to LA). Conversely, three states reported a slightly higher mortality rate in the prison population (CA to AL), and seven states reported a substantively higher mortality rate in the prison population (KS to OH). The SMRs for states with a greater than 50 per 100,000 increase in the prison mortality rate compared to the general population mortality rate were TX 5.55 (95% confidence interval (95CI) 4.56, 6.55), DE 7.51 (95CI 1.95, 13.07), NJ 4.11 (%CI 2.95, 5.28), MI 6.18 (95CI 4.71, 7.64), and OH 10.56 (95CI 8.33, 12.79). The additional age and sex specific data for these calculations come from state sources^14,15,18–25^.

**Figure 2.**
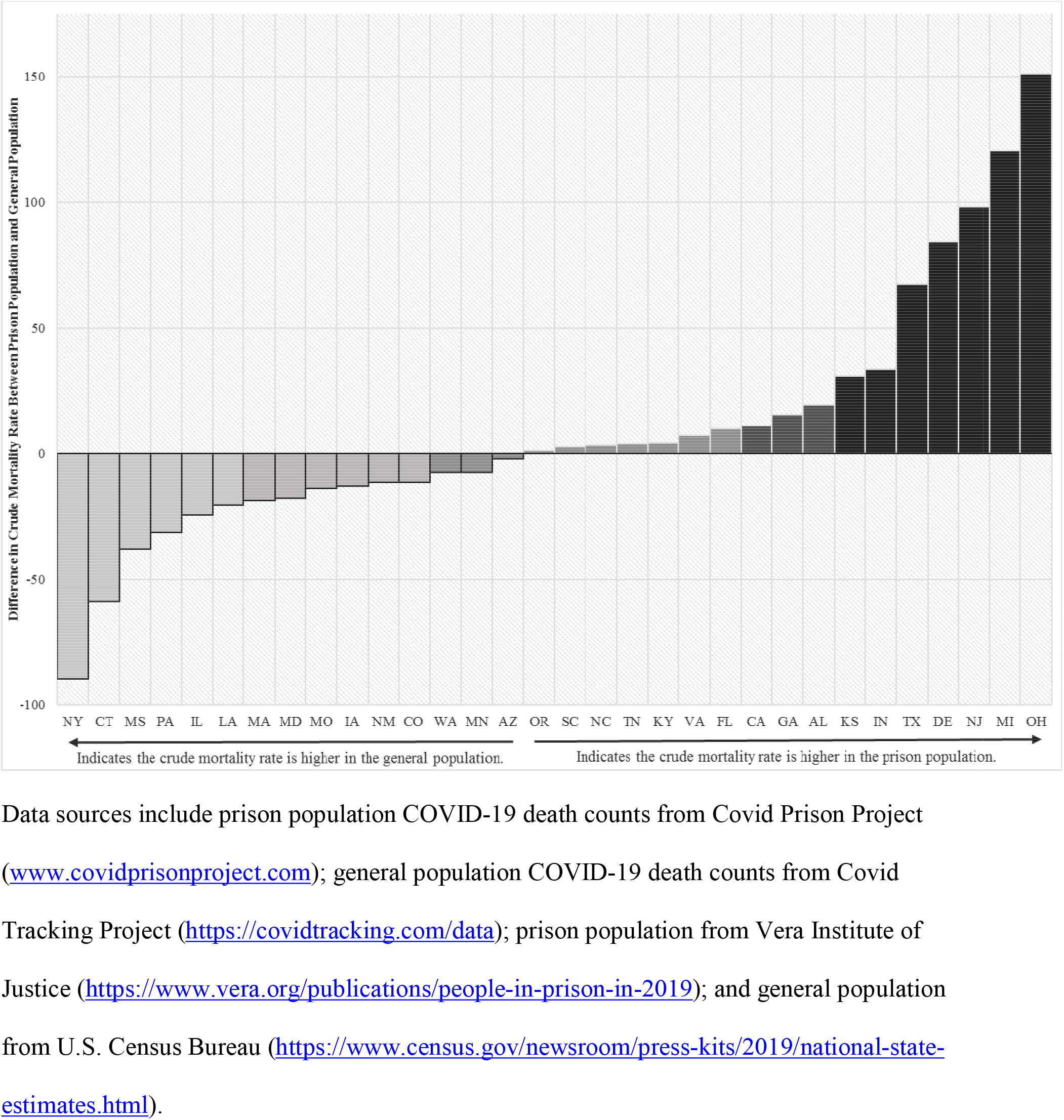
Difference in Crude Mortality Rates (per 100,000) for COVID-19 Related Deaths for the Prison Population compared to the General Population by State, July 15, 2020

Figure 3 shows the number of COVID-19 deaths as a percentage of deaths from all causes reported by departments of correction using the latest year of general mortality data available (2016). DE, MI, NJ, and OH have reported a number of COVID-19 deaths that exceeds 50% of their deaths from all causes. For NJ, the number of COVID-19 deaths in the first 6.5 months of 2020 has almost matched deaths from all causes, potentially indicating that the final all-cause mortality rate for 2020 will be double the rate of previous years.

**Figure 3.**
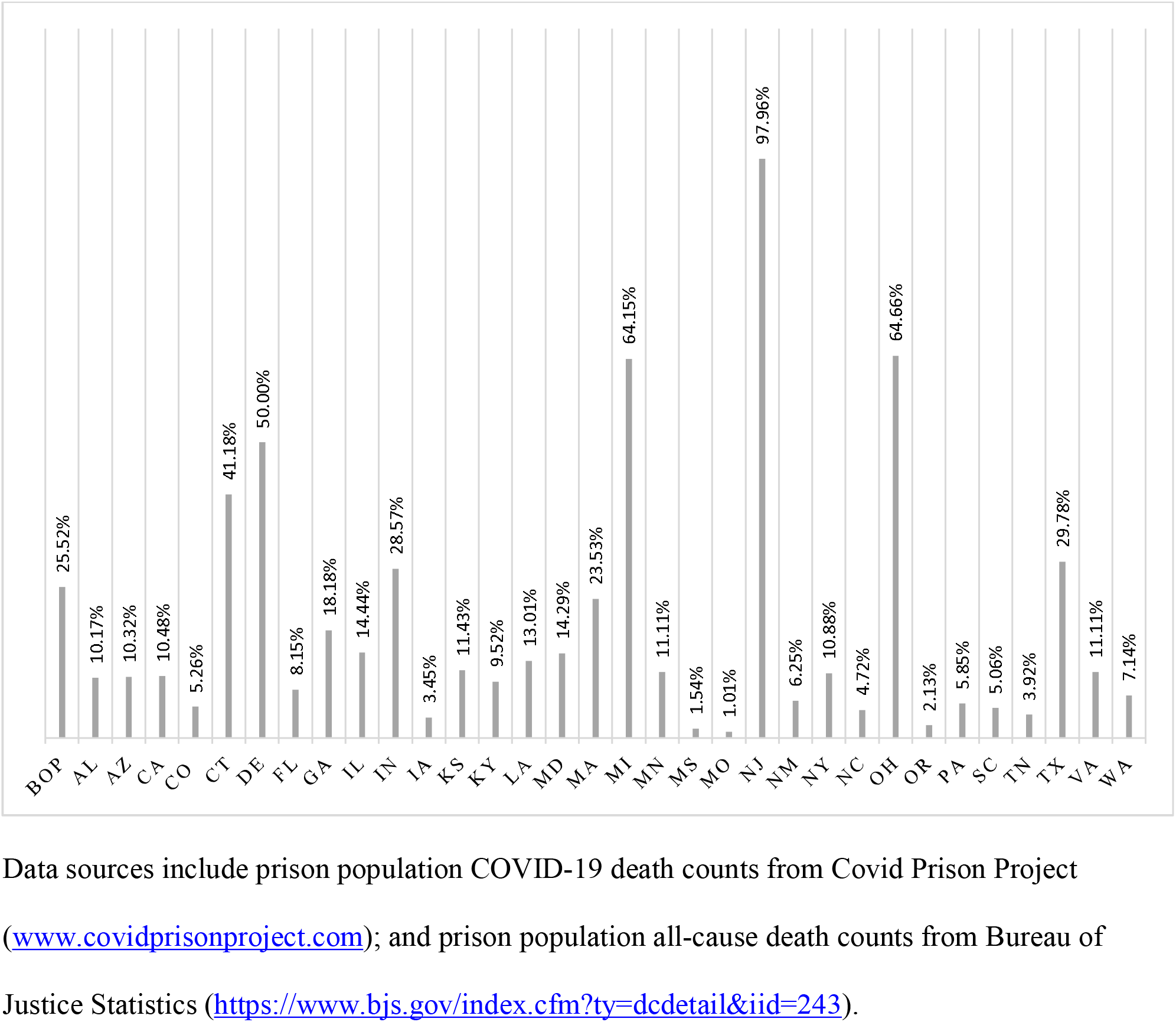
COVID-19 Deaths as a Percentage of Deaths from All Causes in Prison, 2016

Case Study: Texas

The COVID-19 crude mortality rate for prison residents is 79.4 per 100,000, and the corresponding rates for prison staff and the general population are 24.2 and 11.8, respectively. The Texas Department of Criminal Justice (TDCJ) reports information about residents who died in their custody through updates on their website^12^ when an investigation has been completed and COVID-19 has been confirmed to be a contributing cause. As of July 15, investigations for 75 deaths were completed out of 120 total COVID-19 deaths (one case is missing age). Of the 75 completed cases, 45 deaths were among residents aged 65 and older (60.8%), 27 deaths were of residents aged 50 to 64 years (36.5%), and two were of residents below the age of 50 (4.7%). The rates for comparable age groups in the general population of Texas were 73.2%, 20.8%, and 6.0%, respectively. Underlying health conditions were noted in 36 cases. For those cases with reported information (n=41), 10 tested positive for COVID-19 after being hospitalized, 17 were tested the day of hospitalization, and 14 were tested prior to hospitalization. All but three people died in a hospital and the mean length of hospitalization was 9.38 days with a range of less than one day to 30 days. Most deaths that occurred within the custody of TDCJ happened at the Galveston Hospital Unit.

Deaths have been reported in 21 of the 104 units in TDCJ. Three prison units accounted for almost half of all investigated deaths (45.3%). The two units with the largest number of deaths were geriatric facilities. The Duncan Unit housed 0.28% of the prison population in Texas, and was located in a rural county with a population of less than 100,000. This facility had an average age of 65.68 years (SD = 7.91; range 30 to 88 years), a case rate of 66.5%, a case fatality rate of 4.3%, and a crude mortality rate of 29 per 1,000. The case rate among employees at this unit was 36.7%. For comparison, the case fatality rate for the county in which the prison was located was 1.2%, and the case rate was 1.2%. The county had reported 12 COVID-related deaths to date (crude mortality rate was 0.14 per 1,000). A second, larger, geriatric facility, which housed three times as many people and was also located in a small rural county, reported a case rate of 34.5%, a case fatality rate of 2.8%, and a crude mortality rate of 9.7 per 1,000. The average age at this facility was 56.85 years (SD = 12.86, range 23 to 88 years). The case rate among staff was 15.0%. The county in which this prison was located had a case rate of 0.8% and had reported two deaths related to COVID-19 (a case fatality rate of 0.2%, crude mortality rate was 0.02 per 1,000).

## Discussion

This study provides insight into the magnitude of excess deaths in prison settings due to COVID-19 in the U.S. There has been a steady increase in the COVID-19 mortality rate for the prison population overall; however, mortality rates varied by state. Overall, people in prison were experiencing a significantly higher mortality burden compared to the general population (SMR = 2.75). For a handful of states (n = 5), these disparities were more extreme, with SMRs ranging from 5.55 to 10.56. Similarly, four states reported COVID-19 related death counts that are more than 50% of expected deaths from all-causes in a calendar year. The Texas case study found that there was also variation in mortality among units within prison systems, with geriatric facilities potentially at highest risk. Prison facilities are often located in rural counties, and may contribute to the spread of COVID-19 in these areas as staff move in and out of the facility. Indeed, there was some evidence from Texas that COVID-19 case rates were higher among both prison residents and staff compared to the local county.

It is important to note that this analysis does not include deaths that have occurred in other correctional spaces, such as local county jails (e.g., Cook County Jail, IL, Rikers Island, NY), city “lockups” or holding facilities, regional jails, U.S. Marshall facilities, military operated facilities, tribal facilities, or ICE operated facilities. While these are all important correctional spaces that make up the “whole pie” of incarceration^26^, each of these contexts pose unique risks for the transmission of COVID-19. For example, state prisons and BOP house the largest share of people under correctional confinement on any given day (~1.3 million); although, over 10 million people cycle through jails in a given year. Large urban jails such as LA County Jail and Cook County Jail house more people than most single-site prisons. Preliminary analysis from Cook County Jail, IL^27^ shows that jail cycling was associated with 15.7 percent of documented cases in IL, exceeding other known predictors (e.g., population density and public transit utilization). Another important correctional space that is absent from this analysis is community supervision (e.g., probation or parole), which is actually responsible for the largest number of people under correctional supervision (whether in the community or confined to a facility).

Our study is the first to characterize COVID-19 deaths in U.S. prisons, and highlight disparities in mortality between prison residents and people living in the community. Understanding the dynamic trends in COVID-19 mortality in prisons as they move in and out of “hotspot” status^28^ requires a comprehensive understanding of public health, corrections, and justice health^29^. This study also serves as a call to action. We cannot ignore the urgent need to have a focused approach to COVID-19 mitigation in prisons. This will require nuanced analyses of higher quality data both within and across systems. Finally, we would be remiss if we did not mention that, on average, prison systems have only released 5% of their population. Our findings underscore the need for more drastic release efforts such as wider use of compassionate release for geriatric and other vulnerable people^30^ that includes emergency planning for release^31^.

## Data Availability

Data are available from Covid Prison Project.

## References

1. Centers for Disease Control and Prevention. Forecasts of Total Deaths, July 15, 2020. Accessed July 20, 2020: https://www.cdc.gov/coronavirus/2019-ncov/covid-data/forecasting-us.html.

2. Statista. Coronavirus (COVID-19) deaths worldwide per one million population as of July 20, 2020, by country. Accessed July 20, 2020: https://www.statista.com/statistics/1104709/coronavirus-deaths-worldwide-per-million-inhabitants/.

3. Centers for Medicare and Medicaid Services. COVID-19 Nursing Home Data. July 5, 2020. Accessed July 20, 2020: https://data.cms.gov/stories/s/COVID-19-Nursing-Home-Data/bkwz-xpvg/.

4. The New York Times. Coronavirus in the U.S Active Case Clusters. 2020. Accessed July 20, 2020: https://www.nytimes.com/interactive/2020/us/coronavirus-us-cases.html#clusters.

5. Covid Tracking Project. Our Data. Accessed July 16, 2020: https://covidtracking.com/data.

6. Centers for Disease Control and Prevention (CDC). Provisional COVID-19 Death Counts by Sex, Age, and State. Accessed July 16, 2020: https://data.cdc.gov/NCHS/Provisional-COVID-19-Death-Counts-by-Sex-Age-and-S/9bhg-hcku/data.

7. American Community Survey. 2019 National and State Population Estimates. U.S. Census Bureau. Accessed July 16, 2020: https://www.census.gov/newsroom/press-kits/2019/national-state-estimates.html.

8. American Community Survey. 2018 Age and Sex Tables. U.S. Census Bureau. Accessed July 16, 2020: https://data.census.gov/cedsci/table?tid=ACSST5Y2018.S0101&hidePreview=true.

9. Bureau of Justice Statistics. Mortality in Correctional Institutions. U.S. Department of Justice. Accessed July 16, 2020: https://www.bjs.gov/index.cfm?ty=dcdetail&iid=243.

10. Carson AE. Prisoners in 2018. Bureau of Justice Statistics, U.S. Department of Justice. Accessed July 16, 2020: https://www.bjs.gov/index.cfm?ty=pbdetail&iid=6846.

11. Kang-Brown J, Montagnet C, Schattner-Elmaleh E, Hinds O. People in Prison 2019. Vera Institute of Justice. Accessed July 16, 2020: https://www.vera.org/publications/people-in-prison-in-2019.

12. Texas Department of Criminal Justice. Presumed COVID-19 Offender Deaths. Accessed July 16, 2020: https://www.tdcj.texas.gov/covid-19/presumed.html.

13. Texas Department of Criminal Justice. FY 2016 Texas Department of Criminal Justice Statistical Report. Accessed July 16, 2020: https://www.tdcj.texas.gov/documents/Statistical_Report_FY2016.pdf.

14. Texas Department of Criminal Justice. Unit Directory. Accessed July 16, 2020: https://www.tdcj.texas.gov/unit_directory/index.html.

15. Texas Department of State Health Services. News Updates: COVID-19 (new coronavirus). Accessed July 16, 2020: https://dshs.texas.gov/news/updates.shtm

16. Texas Demographic Center. 2018 Estimated Population of Texas, Its Counties, and Places. Accessed July 16, 2020: https://demographics.texas.gov/Data/TPEPP/Estimates/.

17. The Texas Tribune. Texas Prison Inmates. Accessed July 16, 2020: https://www.texastribune.org/library/data/texas-prisons/.

18. State of New Jersey Department of Corrections. Offenders in New Jersey Correctional Institutions on January 1, 2020 by Age. Accessed July 18, 2020: https://www.state.nj.us/corrections/pdf/offender_statistics/2020/2020_Age.pdf.

19. The State of New Jersey. New Jersey COVID-19 Information Hub. Accessed July 18, 2020: https://covid19.nj.gov/.

20. Michigan Department of Corrections. 2018 Statistical Report. Accessed July 18, 2020: https://www.michigan.gov/documents/corrections/MDOC_2018_Statistical_Report_-_2019.07.18_662129_7.pdf.

21. The State of Michigan. Coronavirus. Accessed July 18, 2020: https://www.michigan.gov/coronavirus/0,9753,7-406-98163_98173---,00.html.

22. Ohio Department of Rehabilitation and Correction. January 2019 Census of ODRC Institutional Population, Demographic and Offense Summary. Accessed July 18, 2020: https://drc.ohio.gov/Portals/0/INSTITUTION%20CENSUS_JAN%202019.pdf.

23. Ohio Department of Health. COVID-19 Dashboard. Accessed July 18, 2020: https://coronavirus.ohio.gov/wps/portal/gov/covid-19/dashboards/overview.

24. Delaware Department of Correction. Annual Report, 2019. Accessed July 18, 2020: https://doc.delaware.gov/assets/documents/annual_report/DOC_2019AnnualReport.pdf.

25. State of Delaware. Coronavirus (COVID-19) Data Dashboard. Accessed July 18, 2020: https://myhealthycommunity.dhss.delaware.gov/locations/state#outcomes.

26. Sawyer W, Wagner P. Mass Incarceration: The Whole Pie 2020. Prison Policy Initiative; March 24, 2020, accessed: https://www.prisonpolicy.org/reports/pie2020.html.

27. Reinhart E, Chen D. Incarceration And Its Disseminations: COVID-19 Pandemic Lessons From Chicago’s Cook County Jail. Health Affairs 2020; https://doi.org/10.1377/hlthaff.2020.00652.

28. Hamblett A. Covid Prison Hotspots: July 3, 2020 to July 16, 2020. Covid Prison Project. Accessed July 20, 2020: https://covidprisonproject.com/covid-19-case-watch/.

29. Nowotny KM, Zielinski MJ, Stringer KL, Pugh T, Wu E. et al. Training the Next Generation of Researchers Dedicated to Improving Health Outcomes for Justice-Involved Populations. Am J Pub Health 2020; 110(S1):S18–S20 https://doi.org/10.2105/AJPH.2019.305411.

30. Amend. Requesting Compassionate (Early) Release/ Parole During COVID-19. Amend: Changing Correctional Culture. Accessed June 25, 2020: https://amend.us/requesting-compassionate-early-release-parole-during-covid-19/.

31. Williams B, Ahalt C, Cloud D, Augustine D, Rorvig L, Sears D. Correctional Facilities In The Shadow Of COVID-19: Unique Challenges And Proposed Solutions. Health Affairs Blog March 26, 2020 DOI: 10.1377/hblog20200324.784502.

